# The impact of the variation of imaging factors on the robustness of Computed Tomography Radiomic Features: A review

**DOI:** 10.1101/2020.07.09.20137240

**Authors:** Reza Reiazi, Engy Abbas, Petra Famiyeh, Aria Rezaie, Jennifer Y. Y. Kwan, Tirth Patel, Scott V Bratman, Tony Tadic, Fei-Fei Liu, Benjamin Haibe-Kains

**Author notes:** Author to whom correspondence should be addressed. Benjamin HAIBE-KAINS, PhD. Princess Margaret Cancer Centre, University Health Network, Toronto, Ontario, Canada.

## Abstract

The field of radiomics is at the forefront of personalized medicine. However, there are concerns regarding the robustness of its features against multiple medical imaging parameters and the performance of the predictive models built upon them. Therefore, our review aims to identify *image perturbation factors* (IPF) that most influence the robustness of radiomic features in biomedical research. We also provide insights into the validity and discrepancy of different methodologies applied to investigate the robustness of radiomic features. We selected 527 papers based on the primary criterion that the papers had imaging parameters that affected the reproducibility of radiomic features extracted from computed tomography (CT) images. We compared the reported performance of these parameters along with IPF in the eligible studies. We then proceeded to divide our studies into three groups based on the type of their IPF: (*i*) scanner parameters, (*ii*) acquisition parameters and (*iii*) reconstruction parameters. Our review highlighted that the reconstruction algorithm was the most reproducible factor and *shape* along with *intensity histogram* (IH) were the most robust radiomic features against variation in imaging parameters. This review identified substantial inconsistencies related to the methodology and the reporting style of the reviewed studies such as type of study performed, the metrics used for robustness, the feature extraction techniques, the image perturbation factors, the reporting style and their outcome inclusion. Finally, we hope the IPFs and the methodology inconsistencies identified will aid the scientific community in conducting research in a way that is more reproducible and avoids the pitfalls of previous analyses.

## INTRODUCTION

Computed Tomography (CT) is the modality of choice for the depiction, diagnosis and monitoring of many diseases in the body. Having the ability to provide consistent high-resolution images ushers the way for CT to have extended applicability in medicine such as diagnostic, prognostic, quality assessment (QA), and dose calculation in radiotherapy (Liguori et al. 2015). Moreover, the number of possible applications continues to grow because of the innovative ways researchers have designed to extract new, potentially clinically-relevant features from radiological images (Liu et al. 2019).

Advances in the field of artificial intelligence resulted in introducing decision support based on quantitative image descriptors, as a new tool to assess medical images beyond the narrow visual inspectors. The main idea behind this new research field, called radiomics, is that advanced analysis of images can noninvasively amplify clinical prognostic nomographs, correlate imaging phenotypes with genomic and proteomic signatures, and subsequently reform clinical decision making. Although several challenges remain for bringing radiomics into daily clinical practice, radiomics signatures increasingly become a critical component of precision medicine for the integration of image-driven data towards a more personalized treatment in the near future. (Afshar et al. 2019; Hosny et al. 2018).

As the radiomics field matured, it became apparent that the main drawback of radiomic features is their low reproducibility to variation in acquisition and reconstruction settings which may affect the generalizability of any models built upon those features (Duda, Kretowski, and Bezy-Wendling 2013; Kumar et al. 2012; Zhao et al. 2016; Traverso et al. 2018; Solomon et al. 2016; Yamashita et al. 2019). The effect of variation of image acquisition on the reproducibility of radiomic features has been found greater than that of segmentation (Yamashita et al. 2019) and interobserver variability (Choe et al. 2019). This variation affects the information being extracted by image feature algorithms, which in turn affects classifier performance and is of paramount importance to ensure the successful application of CT-derived radiomics in the field of oncology (Shafiq-Ul-Hassan et al. 2018; Park et al. 2020). Consequently, we must treat the reported performance of radiomics-based models with caution (Mackin et al. 2015) and quantitative changes may be primarily due to acquisition variability rather than from real physio-pathological effects (Andrearczyk, Depeursinge, and Müller 2019). CT-derived radiomic features have intrinsic dependencies on voxel size and number of gray levels, which shows their application is highly dependent on careful selection of the nominal voxel size and the number of intensity bins (Shafiq-Ul-Hassan et al. 2017 (1); Larue et al. 2017; Lee et al. 2019). For instance, the type of binning and number of bins significantly affected radiomic parameters extracted from coronary artery plaques (Kolossváry et al. 2018). In other words, post-processing setting, i.e bin size in intensity normalization or voxel size in voxel rescaling, should be adjusted depending on the type of radiomic feature, imaging factor, organ and clinical outcome. In addition, the numerical values of radiomic features showed to be highly correlated with tumor volume and voxel resampling was not sufficient to remove this correlation (Shafiq-Ul-Hassan et al. 2017). One can eliminate this dependency only by including the number of voxels in feature definitions (i.e., feature normalization) (Shafiq-Ul-Hassan et al. 2018). One possible solution would be to focus on the imaging parameters that affect the robustness of radiomic features. We refer to these non-reproducible imaging factors as imaging perturbation factors (IPFs). Having known the IPFs, we must select the radiomics features robust to them. However, this action requires deep knowledge about these IPFs and their related robust features. In this review, we investigate the IPFs, as well as the least and the most affected radiomic features associated with them. This review aims to provide insights into the validity and the discrepancy of different methodologies applied to investigate the robustness of radiomics features.

## MATERIALS AND METHODS

### Literature Search

We conducted this review from April to July 2020. Reporting complies with the PRISMA-P Preferred Reporting Items for Systematic Reviews and Meta-Analyses statement (Moher et al. 2015). The articles included met all the eligibility criteria given in the subsequent paragraphs. We included only peer-reviewed English full-text reports available in journals that presented full results of reproducibility tests on radiomic features. We intend to have all the articles that match the inclusion criteria, defined in the material and methods section, presented in a statistical analysis of reproducibility study of just CT-based radiomics models. We only included articles that had at least one of the following search words in their titles or abstracts: Radiomics AND (Repeatability OR Reproducibility OR Robustness OR Stability OR Perturbation factor OR Perturbation parameter) specified in their search string. We admitted all PubMed search results up to July 2020.

### Eligibility Criteria and Study Selection

The included articles must include the following eligibility criteria: have radiomics features extracted from CT images, be reproducibility studies, and focus on the reproducibility of image perturbation factors. We excluded 558 studies for reasons such as: being review articles or the studies using other imaging modalities other than CT. Furthermore, we excluded repeatability or test-retest studies because they do not evaluate inter-scanner dependency or the impact of imaging parameters, either of which could affect the results of the study. We also included features or feature sets that were found to be (non)robust in regards to the factors under study. The studies included also had to report at least one or more of the following quantitative outcomes of interest: variability of radiomic features with respect to Image acquisition parameters, scanner type, and Image reconstruction.

We also screened the Cochrane Database of Systematic Reviews for any previous systematic reviews addressing the reproducibility of CT-based radiomic features. For all the articles obtained where we used the full text for data extraction, we screened the bibliographic references within them for potentially eligible studies. The researchers downloaded these electronic full-text articles using university library subscriptions.

### Data Extraction and Synthesis

We extracted information in the studies, including the subject (patient or phantom), the number of cases, the type of organ, the clinical implications for human studies and the type of phantom for phantom studies. We noted the study type (retrospective or prospective) and the model of CT scanner used as well as any image acquisition and reconstruction parameters explicitly stated in the text. We noted the total number of radiomic features tested and grouped these features according to their *Shape* features, *Intensity histogram* (IH), higher-order textural features (*GLCM, RLM, GLSZM, GLDZM, NGTDM, NGLDM*) and their transformation (*LoG, wavelet, Gabor, Sobel, Law*). Complete list of the abbreviations used in this article has been presented in supplementary table 1. We also classified features based on 2D or 3D extraction. Within our classification, we considered features like ROI size (e.g., volume), shape (e.g., eccentricity, compactness), boundary shape (e.g., shape index), sharpness (e.g., sigmoid slope) as shape features. In addition, we also recorded the details of the software used to quantitatively extract radiomic features, as well as the statistical methods used. We extracted information regarding the type of metric used to report feature robustness. Three main metrics are intra-class correlation coefficient (ICC), concordance correlation coefficient (CCC), and Coefficient of Variation (COV). COV is the ratio of the standard deviation to the mean. ICC and CCC are very similar metrics. ICC evaluates the clustering of features from several classes using the correlation of features within classes and CCC measures the agreement between two variables.

**Table 1.** Abbreviation

| Term | Definition |
| --- | --- |
| ACQ | Acquisition |
| CCC | Concordance Correlation Coefficient |
| CCR | Credence Cartridge Radiomics phantom |
| COV | Coefficient of variation: $\text{abs}(\text{SD}/\text{mean}) \times 100\%$ |
| FD | fractal dimension analysis |
| FOV | field of view |
| GLCM | Gray Level Cooccurrence Matrix |
| GLDZM | gray-level distance zone matrix |
| GLSZM | Gray level Size zone Matrix |
| ICC | intraclass correlation coefficients |
| IH | Intensity Histogram |
| LBP | local binary patterns |
| LoG | Laplacian of Gaussian |
| LMEM | linear mixed-effects model |
| NGLDM | neighboring gray-level dependence matrix |
| NGTDM | Neighborhood Gray tone difference matrix |
| REC | Reconstruction |
| RLM | Run Length Matrix |
| SCA | Scanner |
| UWP | uniform water phantom |

## RESULTS

The PubMed search yielded 597 abstracts, including 38 eligible studies that reported the effect of imaging parameters on the reproducibility of radiomic features extracted from CT images and/or features or feature sets found (ir)reproducible to the factors under study. We retrieved the full text for 42 abstracts deemed suitable for in-depth evaluation, including 4 located in the references of retrieved studies (Figure 1). After full-text evaluation, 7 studies were further excluded because, despite their title and abstract, they did not meet the inclusion criteria. We derived a qualitative synthesis from 35 studies, of which 20 studies used retrospective human data, and 15 used prospective phantom data.

**Figure 1.**
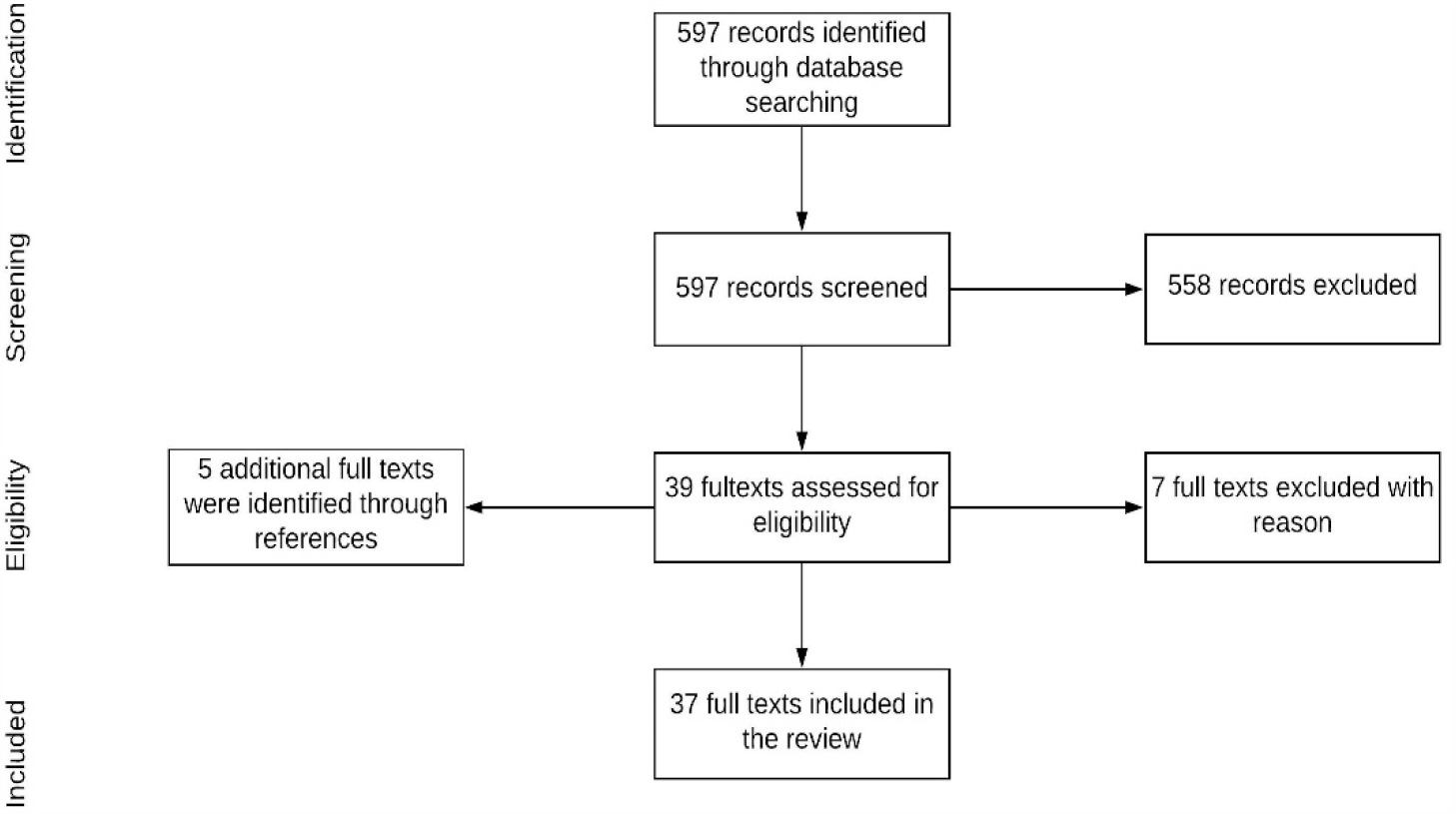
The PubMed search yielded 597 abstracts for screening against our selection criteria, reduced to 38 eligible for full text. The full text was retrieved for 37 abstracts deemed suitable for in-depth evaluation, including 5 that were located in the references of retrieved studies

The vast majority of retrospective human studies addressed the reproducibility of CT-based radiomic features in cancers such as lung (76%), liver (12%), head and neck (4%), rectal (4%), and pancreatic (4%) (Figure 2A). The number of patients reported in the retrieved studies ranged from 10 to 104, however on average, the studies included 40 patients. Amongst these studies, only one used cone-beam CT (CBCT); all others used planning or diagnostic CTs. All prospective studies used different types of physical radiomic phantoms. Amongst the 15 prospective phantom studies, Credence Cartridge Radiomics (CCR) Phantom developed by Mackin (Mackin et al. 2015) was the most common phantom used (58%) followed by Anthropomorphic (30%), CT texture analysis (CTTA) (6%) and water phantom (wp) (6%) (Figure 2B).

**Figure 2.**
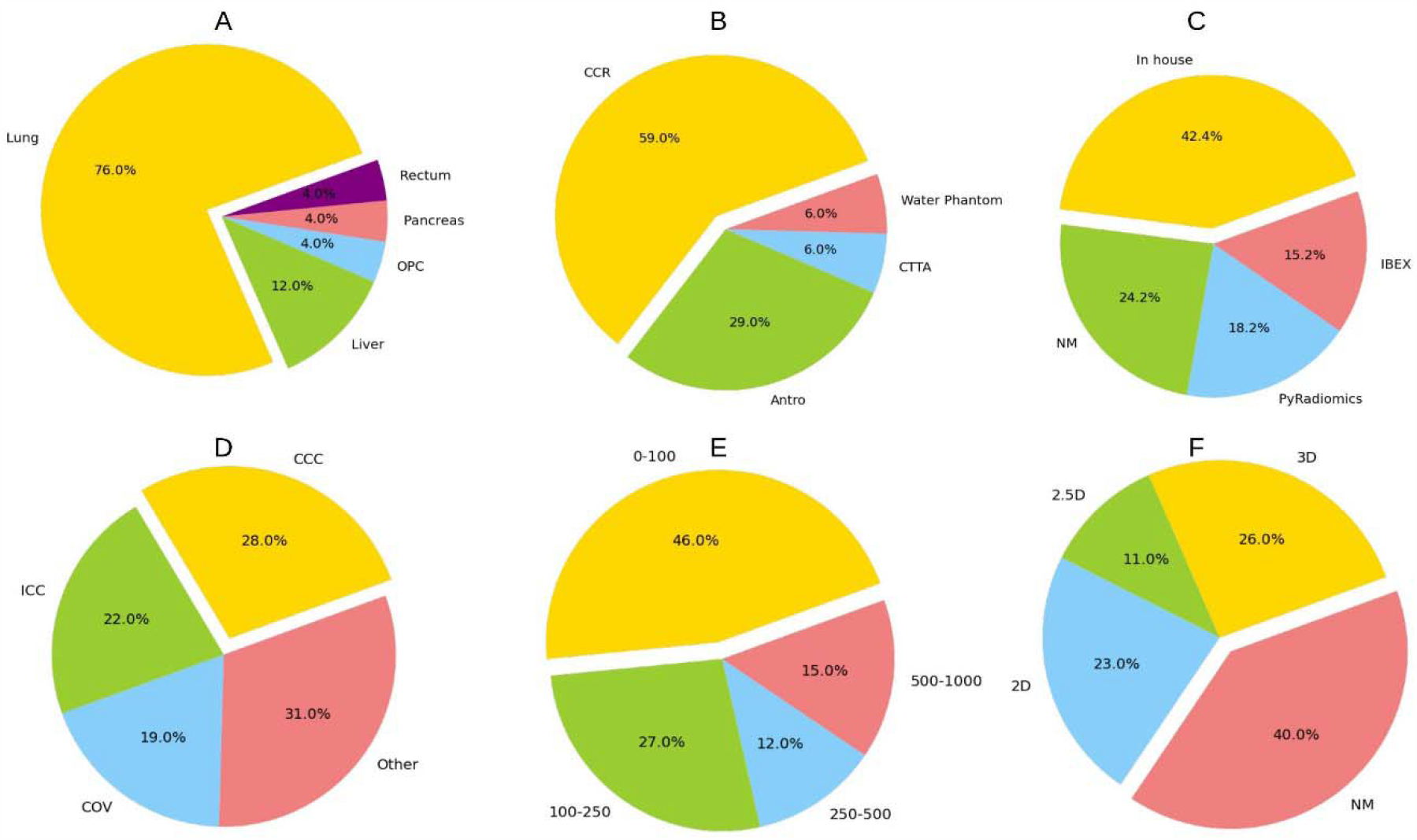
Reviewed studies based on type of organ studied in human studied (A), common phantom type in phantom studies (B), software used for feature extraction (C), metric used to report features reproducibility(D), distribution of the total number of features extracted (E) and feature dimensions (F)

### Image perturbation factors

We grouped the Image perturbation factors (IPF) into 3 classes: (i) scanner; (ii) acquisition (including patient-related parameters); and (iii) reconstruction (Supplementary Table 2). The frequency of reporting a specific imaging parameter was depicted in figure 3. The scanner studies are those studies that only focused on the effect of different CT scanners disregarding any specific acquisition or reconstruction parameter. Out of the selected studies, 17 publications investigated the effect of reconstruction factors alone or in combination with acquisition parameters and manufacturer dependency (Supplementary Table 3). We then evaluated the acquisition parameters in 4 of the studies. Similarly, another 3 studies evaluated scanner dependency (Table 3). None of these studies were prospective because of concerns over repeated patient exposure to radiation without expected therapeutic benefits.

**Table 2.** Information extraction table

| *Type of Data* | *Extracted information* |
| --- | --- |
| **General** | Type of imaging factor (Acquisition, Reconstruction or Scanner), Scope (Retrospective \| Prospective), Type of study (Repeatability or Reproducibility) |
| **Data** | Phantom (type) or Patient (Organ), Number of cases |
| **Imaging** | **Acquisition:** Scanner Model, Scan Type, Det Col, mAs, mA, KvP, Dose, Eff.Tube Current, Pitch, Focal spot size, s<br>**Reconstruction:** Recon Dia & pixel Size (FOV), Recon Algo & Conv Ker, SNR/noise, WW/WL,<br>**Patient protocol,:** Injection Rate, Patient position, Breathing or respiratory motion, Contrast dose, scatter(weight/size), type of perfusion |
| **Analysis** | **Feature extraction:** Software, language, Feature Dimension, Total number of features,type post processing<br>**Reproducibility index:** Type and Cut off<br>**Outcome prediction:** Type |
| **Results** | number of stable features, most and least reproducible feature or feature set, most and least variable perturbation factor |

**Table 3.** Summary of selected studies to evaluate the clinical outcome of radiomic feature robustness

|  | Obj | SC | # P | SO | Metric | O.C | FUI | # | RFe | LRF | MRF | D.M | ROI |
| --- | --- | --- | --- | --- | --- | --- | --- | --- | --- | --- | --- | --- | --- |
| (Berenguer et al. 2018) | CCR | Pros | na | IBEX | COV<0.1 | No | mAs, FOV,kVp, Rec.Ker | 177 | shape & IH | kVp, Rec.Ker | Pitch | y | Seg |
| (Shafiq-ul-Hassan et al. 2017) | CCR | Pros | na | In.h | COV<0.1 | No | mAs, Pitch, Rec. Ker | 88 | GLSZM | Reco. Ker | mAs, Pitch | y | Sph |
| (Varghese et al. 2019) | CTTA | Pros | na | NM | PAD<0.15 | No | Slice.Th, FOV, mA, kVP | 235 | Entropy of FFT mag & ph | Slice.Thi | FOV | y | Sph |
| (Y. J. Kim et al. 2019) | Antro | Pros | na | In.h | MRA,p-value>0.05 | No | Slice.Th, Rec.Ker, mAs | 20 | - | Slice.Th, Rec.Ker, mAs | - | n | Seg |
| (Larue et al. 2017) | CCR | Pros | na | In.h | CCC>0.85 | No | mA, Slice.Th | 114 | GLSZM (SAE) | Slice.Th | mA | y | Sph |
| (Lo et al. 2016) | WP+Lung nodule | Pros | 33 | NM | Q <1 | No | mAs, Pitch, Rec.Ker, Rec,Alg, Dose | 34 | IH mean | Dose | Rec. Alg | y | Sph |
| (Mackin et al. 2018) | CCR | Pros | na | IBEX | COV | No | mAs | 48 | IH | - | mAs | y | rect |
| (Midya et al. 2018) | Antro | Pros | na | In.h | CCC>0.9 | No | mA, Rec.Alg | 248 | GLCM-sum-Ave | mAs | Rec. Alg | n | Seg |
| (Perrin et al. 2018) | Liver | Retr | 38 | In.h | CCC>0.9 | No | FOV(Pixel Spacing) | 254 | (5) LBP | FOV(Pixel Spacing) | - | y | Seg |
| (Yamashita et al. 2019) | Pancreas | Retr | 37 | In.h | ICC>0.8 | No | mAs | 266 | RLM nonuniformity | - | mAs | y | Seg |
| (Solomon et al. 2016) | Lung,Liver,Kidney | Retr | 72 | In.h | ICC | No | Rec.Alg, Dose | 23 | z-axis extent | Rec.Alg, Dose | - | - | Seg |
| (Duda, Kretowski, and Bezy-Wendling 2013) | Liver | Retro | 29 | - | COV | cirrhotic and healthy liver | Slice.Th | 155 | Autocorr | - | Slice. Thk* | n | Seg |
| (Choe et al. 2019) | Lung Nodule | Retr | 104 | PyR | CCC>0.85 | No | Con.Ker | 702 | IH_skewness | Con.Ker | - | n | Seg |
| (Kolossváry et al. 2018) | Coronary Artery plaques | Retr | 60 | Pictologic | ICC>0.8 | No | Rec.Alg | 164 | - | - | Rec. Alg | n | Seg |
| (Larue et al. 2017) | CCR | Pros | na | In.h | Max.Difference | No | Slice.Th, mAs | 114 | GLSZM – Small Area Emphasis (SAE) | - | - | n | Sph |
| (Li et al. 2018) | Lung Ad Ca | Retro | 51 | In.h | CCC>0.8 | EGFR) mutation | Slice.Th, Con.Ker | 1695 | LoG_Entropy_Sigma2.5_2D | Slic.Th | Con. Ker | n | Seg |
| (L. Lu et al. 2016) | Lung | Retro | 32 | NM | CCC>0.8 | No | Slice.Th, Con.Ker | 89 | spherical cap | Conv.Ker | Slic.Th | n | Seg |
| (Zhao et al. 2016) | Lung | Retro |  | NM | CCC>0.85 | No | Slice.Th, Rec.Ker | 89 | Volume | Slice.Th | Conv. Ker | n | Seg |

**Figure 3.**
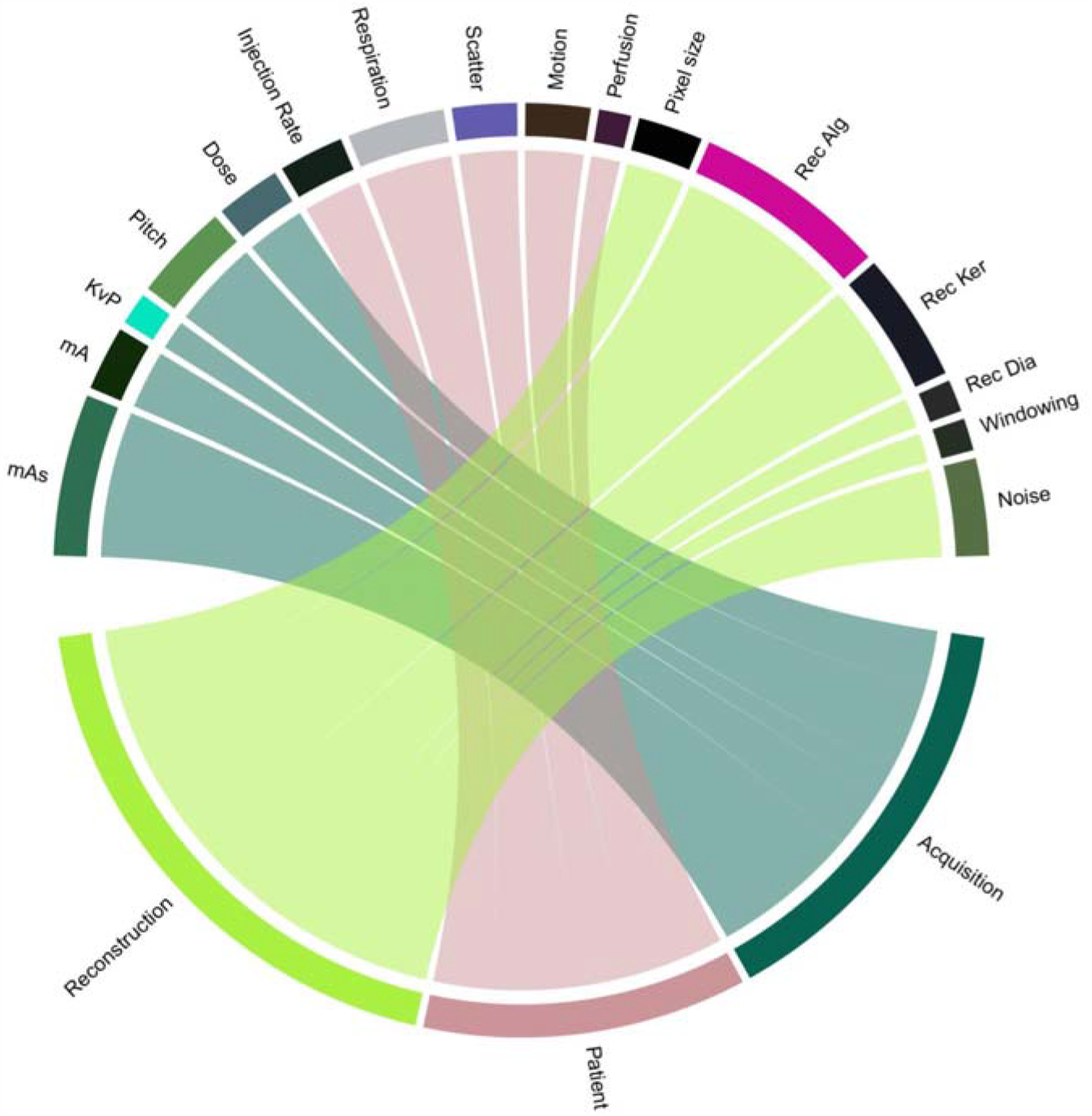
Distribution of image perturbation factors have been studied. Thickness of edges is showing the frequency of each parameter studied. Abbreviations are explained in supplementary

The studies reported some features individually against specific imaging parameters: *GLSZM-SAE* (slice thickness) (Larue et al. 2017), *GLRLM-SRE* (respiration) (Lafata et al. 2018), GLCM-dissimilarity (slice thickness), *IH-Kurtosis, LGRE(mAs), IH-Correlation* (reconstruction kernel) (Midya et al. 2018), gray□level *GLRLM-nonuniformity*(motion), *IH-skewness*(Scatter) (Fave et al. 2015), GLCM-sumAverage(slice thickness) (Duda, Kretowski, and Bezy-Wendling 2013). A short list of recommended features deemed robust against the imaging parameter under investigation is reported on the supplementary table 4. Among the imaging factors, reconstruction algorithm (5 out of 7 papers) and slice thickness (3 out of 5 papers) were both reported to be mostly reproducible against the other factors under study. Pitch and mAs were each reported once to be both reproducible and non-reproducible. Within the reported factors the majority were non-reproducible most of the time (reconstruction kernel) or at least once (KVp, noise, respiration, FOV, windowing, injection, and pixel size) in the reviewed studies. The following sections discuss details of each groups of IPF separately:

**Table 4.** List of recommended radiomic features

| Reference | Recommended features |
| --- | --- |
| (Zhao et al. 2016) | GTDM_Strength, Sigmoid-Offset, GLCM_Sum-Average, GLCM_Sum-Variance, LoG_MGI-s4, LoG_Entropy-s4, Sigmoid-Offset, maximal diameter, maximal diameter and its maximal perpendicular diameter, Volume |
| (Lu et al. 2016) | spherical cap, Sigmoid-Offset-mean, LoG_Entropy_Sigma2.5, IH mean, GLCM_sum-avg, LoG_mean_Sigma2.5 |
|  | GLCM_invDiffnorm.Stats_median.GLSZM_LAE.GLDZM_LILDE.NGLDM_DV.GLCM_maxCorr.GLSZM_SZV107.Stats_mean.NGTDensity_MeanDM_busyness.NGTDM_strength.Stats_skewness.Stats_kurtosis.GLSZM_HILAE.GLSZM_SZ |
| (Kolossváry et al. 2018) | All except for: first-order parameters: mode, harmonic mean and minimum and GLCM parameters: inverse difference sum and sum variance |
| (Duda et al. 2013) | Autocorr, FractalDim, GLCM SumAvg, IH Avg, RLM (ShortEmp, LongEmp, Fraction, HighGLREmp, RLEntr), Laws Texture Energy (E3L3, S3L3, S3E3, E3E3, S3S3), GLDM (DAvg, DEntr, DAngSecMom, DInvDiffMom) |

**Scanner** dependency was evaluated only on two phantom studies. In these studies, the researchers used CCR phantom images from different scanners by different manufacturers with various tube voltages, tube currents (Yasaka et al. 2017), pixel spacing, pitch thickness, slice thickness, reconstruction kernel and dose level (Mackin et al. 2015) to mimic routine imaging protocols. These studies used different feature extraction software (IBEX (Zhang et al. 2015) vs. TexRAD™(Feedback plc, Cambridge, UK)), evaluation metrics (ICC vs mean and SD of the HU) and feature sets (first-order statistics alone vs first-order statistics plus *NGTDM* texture features). They concluded that they should consider scanner dependency since most features have significant scanner dependency and they have to consider and minimize their effects in future radiomics studies. The variation of these features in the phantom images are due to fundamental design differences of the scanners and/or differences in acquisition parameters. It is noteworthy to mention that different CT scanners have been proven to have variation in their Hounsfield units even with the same acquisition parameters (Varghese et al. 2019; Shafiq-ul-Hassan et al. 2017). Perrin et al. showed that utilizing images from different scanners reduced the number of liver tumor-derived robust features (CCC>0.9) from 75 to 35 (out of 254) (Perrin et al. 2018). However, Mackin et al. reported that variations of radiomic features among different scanners were found to be similar to their variation among 20 NSCLC patients stating Busyness-NGTDM and strength-NGTDM the most and least robust features respectively (Mackin et al. 2015). NGTDM textural features reflected the intensity differences between a voxel and its neighboring voxels (Amadasun and King 1989).

**Acquisition** parameters include scan type (helical vs axial), slice thickness, exposure time, exposure (mAs), tube current (mA), tube voltage (kVp), pitch, the field of view (FOV), dose index and focal spot size. In 11 studies researchers investigated one or more of these factors. Amongst these 11 studies, 9 used phantoms and 2 used retrospective human datasets to evaluate dose (Solomon et al. 2016) and slice thickness (Duda, Kretowski, and Bezy-Wendling 2013). The focus of the latter study was on the influence of different liver perfusion imaging (arterial vs. portal). Amongst these imaging parameters, the least problematic imaging factors stated were pitch (Berenguer et al. 2018) once and mAs (Mackin et al. 2018; Lo et al. 2016; Shafiq-ul-Hassan et al. 2017; Midya et al. 2018) four times. In contrast, FOV (Varghese et al. 2019), pitch (Shafiq-ul-Hassan et al. 2017), and the windowing (Y. J. Kim et al. 2019) have all been reported as low reproducible factors (Figure 4).

**Figure 4.**
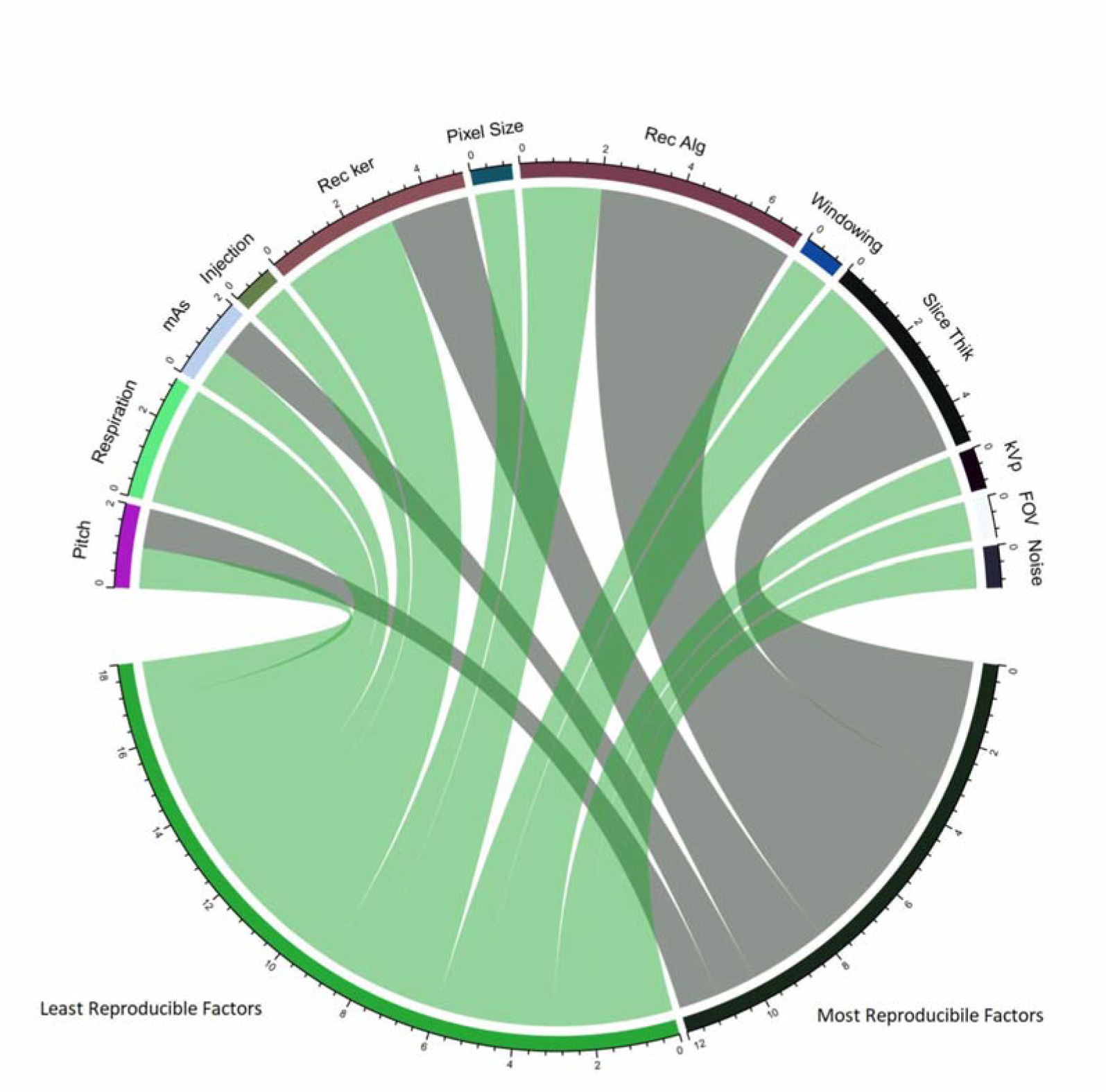
Distribution of most and least reproducible factors reported. Number on the scale shows the quantity of paper reported as the adjacent factor as most or least reproducible factor against the other factors studied.

**Patient-related** parameters include the influence of the scatter as a result of the patient’s weight (size), contrast dose, injection rate, respiration, and type of perfusion. Amongst those factors, we found respiratory motion studied in three studies and contrast injection rate along with patient-related scatter investigated each in only one study (Figure 2). In some papers, they did not state the effect of respiratory motion on feature robustness explicitly, but the superiority of using average intensity projection (AIP) images for radiomic studies over those taken with free-breathing (FB) has been shown (Huynh et al. 2017). Using scans at the end of the exhalation (EoE) phase of a 4DCT acquisition (Choe et al. 2019) has been shown to yield more robust radiomics features (CCC > 0.85). Another study stated that the considerable effect of respiration on feature robustness and they suggested applying 4D stability as a feature selection to reduce feature variability (Du et al. 2019). The reviewed studies reported Shape features (Huynh et al. 2017; Du et al. 2019) and Short-Run Emphasis (Lafata et al. 2018) as the Radiomic features most robust to respiration. Although 4D features were more robust, there was no significant correlation between feature robustness and prognostic value (Figure 4). In another study, contrast enhancement in the delayed phase of CT images for NSCLC patients affected some of the radiomic features and the variability of radiomic features due to contrast uptake was found to be dependent largely on the patient characteristics (Kakino et al. 2020).

**Reconstruction** parameters include reconstruction algorithm, reconstruction (convolution) kernel, reconstruction diameter and pixel size. Fourteen studies — alone or in combination with other factors — focused on the influence of reconstruction parameters (Figure 2). Reconstruction algorithm (7 papers), reconstruction kernel (6 papers) and slice thickness (6 papers) were the most frequent factors which were studied in this group (Table 3). Two studies investigated the effect on the noise index resulting from reconstruction algorithms (H. Kim et al. 2016; Midya et al. 2018). One paper studied the effect of intensity windowing (H. Lu et al. 2019). Reconstruction algorithms were the most robust parameter in five papers (Solomon et al. 2016; Zhao et al. 2016; Midya et al. 2018; Lo et al. 2016; Kolossváry et al. 2018) and the least reproducible imaging parameters in two others (Berenguer et al. 2018; H. Kim et al. 2016). Another parameter reported was the reconstruction kernel which was the most reproducible parameter in two papers (Shafiq-Ul-Hassan M(2), Li Yajun) and the least reproducible parameter in another three papers (Choe J, Kim Y, Lu L).

### Variation in methods

Selected studies varied significantly in terms of the software used for feature extraction, the number of extracted features, and the metrics used for reporting reproducibility. The distribution of studies according to the above discrepancies are as follows:

**Software** details (application framework used for analysis, programming language, and version) were not reported in the majority of studies (Figure 2C). The majority of studies described their software as in-house software mostly based on MATLAB® (The Mathworks®, Natick, MA) (“MathWorks - Makers of MATLAB and Simulink” n.d.). PyRadiomics (Python) (van Griethuysen et al. 2017) and IBEX (Matlab) (Zhang et al. 2015) were the second and third most used feature extraction software respectively.

**The total number of features** extracted was different in these studies. The range of the total number of features was from 5 to 1,695 and the majority fell between 5-100 (45%) (Figure 2E). *Intensity histogram, Gray level co-occurrence matrix, Run-length Matrix* and *Shape* features were the most common feature types used. One ambiguity in the feature calculations was the feature dimension. The majority of studies (40%) did not mention the dimension of feature extraction. However, in 25.7%, 22.9%, and 11.4% of studies that stated their features they used 3D, 2D and 2.5D (averaging 2D features over the slices that cover the segmented ROI instead of real 3D calculation) features respectively (Figure 2F).

**Reproducibility metrics** strongly differed across studies. The metrics encountered were the CCC in 27.8% of studies, ICC in 22.2% of studies and COV in 19.4% of studies. 30% of studies used other metrics such as variability index, interquartile range, proportional difference, absolute difference, or linear mixed-effects models. Some studies reported more than a single metric (Brenquer). However, the specific cut off values used to segregate stable from unstable features were not always stated and some studies just reported the percentage of robust features with different cut off values regardless of which cut off value was suitable. Among the 14 studies using the CCC or (and) ICC metric, the most common cut off values were 0.8 (6), 0.9 (5), and 0.85 (3). For COV, cut off values included 1%, 5%, 10% and 20%. Also, the clustering-based metrics have some limited usage such as the Gaussian mixture model (GMM) homogeneity, completeness, and the V-measure (Andrearczyk, Depeursinge, and Müller 2019).

**Clinical outcome** of feature robustness was only investigated in 2 studies. In those studies, we assessed the influence of the robust features on the model’s performance while applying different imaging parameters. In (Li et al. 2018) the model trained over uniform imaging parameters to predict EGFR mutation in lung adenocarcinoma patients. This study concluded that an imaging dataset with uniform imaging parameters is considerably more efficient than building a model upon robust features extracted from a dataset with non-uniform imaging parameters. Also, (Duda, Kretowski, and Bezy-Wendling 2013) showed that models trained and tested on the same slice thickness had better accuracy over the models that they had trained and tested over data with heterogeneous slice thickness or even trained with one slice thickness and tested over data with different slice thickness. That study recommended that any research concerning the robustness of radiomics features should be followed by outcome prediction. This will help to investigate how useful the robust features are in that specific outcome prediction. In another study multi-window CT based radiomics features out-performed single CT window settings to predict growth patterns of lung cancers (H. Lu et al. 2019). These results confirmed that applying different CT windowing (i.e intensity clipping) would result in selecting different radiomic features which may have different prediction efficacies.

## DISCUSSION

This review investigates the robustness of hand-engineered radiomic features in the context of varying imaging parameters through a deep-dive literature review. In order to make this feasible, imaging parameters, which affect the feature robustness, were also collected and grouped into the most and the least reproducible parameters.

Overall, the number of studies addressing the effects of CT imaging factors on feature robustness is low with the majority of them being phantom studies or retrospective human studies. Although phantom studies are a good guide to conduct prospective studies, many feature values are different between human tissue and phantom material. Even features that we acquired under similar acquisition conditions (such as uniform water phantom) are different from human tissues (Lo et al. 2016). Moreover, there is a lack of comprehensive prospective human studies. This is particularly true when it is in regards to investigating the effects of image acquisition parameters. This is due to our inability to rescan patients in absence of a clinical indication. The differences in the scan parameters have significant effects on almost all radiomics features in all studies. Nevertheless, *Shape* (Berenguer et al. 2018; Yasaka et al. 2017; H. Kim et al. 2016; Zhao et al. 2016; L. Lu et al. 2016) and *FO* (Berenguer et al. 2018; Lo et al. 2016; Mackin et al. 2018; Choe et al. 2019) features were the most robust feature classes across the reviewed papers. This is logical because features rely on the segmented tumor boundaries and/or because features have low-frequency change components, such as shape. They are also less dependent on the imaging parameters than the feature groups that are characterized by the high-frequency change components like texture. In one study, *GLRLM* was found to be robust against the reconstruction algorithm (Kolossváry et al. 2018). Previous studies have shown that pre-processing reduces the effect of image parameter variation on feature robustness (Larue et al. 2017; Shafiq-Ul-Hassan et al. 2018). However, there is no universal agreement about the type and the details of the preprocessing setting. In a study to evaluate the radiomic feature stability in lung cancer, the researchers found that down-sampling small voxels to large voxels, and thus, creating simple averaging, the process became more desirable compared to up-sampling large voxels to small voxels. The rationale behind this is due to the process potentially involving interpolation with bias (Lee et al. 2019). These findings suggest that post-processing could affect reproducibility. They also suggest more precise techniques may be required to ensure optimal pre-processing settings. As shown in previous sections, the robustness of radiomic features depends on the imaging parameters as well. It means global pre-processing settings would not be effective in removing dependencies for all imaging parameters. Although imaging parameters will remain the main reason for feature robustness, there are other unseen factors which make this even more complex. One of these factors is the intrinsic intra-patient variation of radiomics features. It was shown that the number of robust features between normal tissue-derived and tumor-derived features within the same patient is different even with the same imaging parameters (Yamashita et al. 2019; Perrin et al. 2018). The other interesting factor is patient age, which is worth paying more attention to, for future studies to evaluate the severity of the mentioned factors (Boughdad et al. 2018). Lastly, the sensitivity of radiomic features to imaging parameters was shown to be inherently organ dependent (Mahon, Hugo, and Weiss 2019; Mackin et al. 2018; Fave et al. 2015; Berenguer et al. 2018) or patient-specific (Kakino et al. 2020).

The main objective of this review was to collect a list of robust features and the most reproducible imaging factors. We assumed that the resulting list would help direct future radiomics studies. However, this review came short on that front due to the substantial inconsistencies related to the methodology and the reporting style of the reviewed studies. We summarized some of these discrepancies as follows:

### Type of studies

The prospective studies were mostly conducted using phantoms. Phantom-based studies have been done to remove the need for additional exposure to radiation to the patients. However, the applicability of these studies is limited because of the inherent significant difference between phantom material and human tissue. Most human studies were retrospective with a select few prospective patient studies done on reconstruction parameters. The drawbacks of the retrospective studies are that the investigators did not have control over the factors studied and the scan acquisition parameter variations range were limited to those used in imaging patients. Presently, studying the effects of image acquisition parameters in a prospective patient study is not feasible. It is without a doubt that prospective studies would be most optimal to evaluate radiomic feature reproducibility and repeatability.

### Metric for robustness

We found the current literature varied with regards to the optimal metric to use for analysis. Nevertheless, There was no statistically significant difference when using either the CCC or the ICC metric (Hu et al. 2016). However, Brenquer found that the choice of metric as well as the threshold influenced the results. Thus, one should give careful attention when choosing a metric and a threshold (Berenguer et al. 2018).

### Feature extraction

Across the papers analyzed for this study, there was heterogeneity in the total number of features calculated, the type of features, and the feature dimensions. In addition, they used a wide variety of software for image processing and feature extraction. Different software platforms have also shown a significant effect on the statistical variation of Radiomic features (Fornacon-Wood et al. 2020). The other issue regarding feature extraction is the feature dimensions. Zhao et al. compared a shape feature and the histogram-derived density statistical features computed from 2D and 3D images and found that the 3D features were more robust than the 2D features across all imaging settings (Zhao et al. 2016). This issue has been stated in another study (Ng et al. 2013).

### Imaging Factor

The studies in this review did not cover all the available and effective imaging factors such as focal spot, scan type, patient positioning, etc. They had also selected different ranges of values for the same imaging factor in different studies. Another issue is that different scanners do not use the same acquisition settings and therefore it is not possible to compare some parameters such as dose and noise index against one another. Therefore, having the unique index to unify the setting like CTDI among different scanners would be desired. Unfortunately, this approach is not practical either. Even with similar acquisition protocols, different scanner types can influence radiomic feature values. Features were significantly affected by noise-related parameters such as slice thickness, the type of reconstruction algorithm (FBP vs. iterative) and patient thickness. This highlights the importance of the noise level.

### Reporting

Most studies investigated the effects of a combination of factors while in reality all other factors should be kept constant except for the one under study. One perfect example of this was the study by (Berenguer et al. 2018). Other problems with the reporting of the investigated studies are the inconsistencies in reproducibility, the percent of robust features, the robust features against all the imaging parameters and the robust feature-factors that determine which features are robust against which factors.

### Outcome inclusion

The lack of clinically labelled human imaging datasets was another shortfall of the available studies. In Duda et al., the feature stability, expressed by its coefficient of variation, was not considerably influenced by the slice thickness (Duda, Kretowski, and Bezy-Wendling 2013). However, in that same study, the classification of the cirrhotic liver using only robust features that were trained and tested on different slice thicknesses resulted in an accuracy fall of 90 to 62. This shows the importance of this issue since any conclusions that were made about feature reproducibility without investigating the effect of the clinical prediction would therefore not be reliable.

## CONCLUSION

Radiomic features stand to play an important role in guiding personalized cancer treatment. However, the robustness of these features against variation in medical imaging parameters may affect the performance of the models built upon these features. Although the reconstruction algorithm was reported to be reproducible more than other reviewed IPFs, there is not enough evidence to support this idea as a result of the methodological inconsistencies which were reported in the results. We also found *shape* as well as IH features to be the most robust radiomic features in some specific cases.

Furthermore, we suggest reducing the dependency of radiomic features on scanner parameters as a key step in developing radiomic models. Two desirable but less practical solutions of doing this are (*i*) credentialing CT scanners used in radiomics studies or correcting for the parameters of the scanner during data analysis; and (*ii*) adopting standardized image acquisition and reconstruction factors. Although pre-processing settings vary depending on the type of features and image acquisition parameters, the most practical and desirable solution would still be implementation of pre-processing procedures, which are not feature (and patient) specific. Although currently there are no universal agreements on type and details of resampling, perhaps a future step for the community would be determining specific pre-processing settings. Our review also recognizes that there are other factors that could affect reproducibility other than imaging parameters including intrinsic intra-patient variation, patient’s age and the organ-specific sensitivity of radiomic features. These are potential, confounding factors that future reproducibility studies should account for.

Finally, we concluded that there is a large discrepancy in methodology among the reviewed studies from the software used to the clinical outcomes investigated. A clear take away from our review was the need for further comprehensive studies like the one that has been done in (Meyer et al. 2019) or (Berenguer et al. 2018) to further investigate the effects of imaging parameters. We would also suggest establishing a universal methodology and reporting styles when it comes to imaging parameters studies that would make the researchers work more reproducible and create higher consistency within the scientific community.

## Data Availability

This is a review article and all data is available online.

## SUPPLEMENTARY TABLES

## REFERENCES

1. Afshar, P., A. Mohammadi, K. N. Plataniotis, A. Oikonomou, and H. Benali. 2019. “From Handcrafted to Deep-Learning-Based Cancer Radiomics: Challenges and Opportunities.” IEEE Signal Processing Magazine 36 (4): 132–60.

2. Amadasun, M., and R. King. 1989. “Textural Features Corresponding to Textural Properties.” IEEE Transactions on Systems, Man, and Cybernetics 19 (5): 1264–74.

3. Andrearczyk, Vincent, Adrien Depeursinge, and Henning Müller. 2019. “Neural Network Training for Cross-Protocol Radiomic Feature Standardization in Computed Tomography.” Journal of Medical Imaging (Bellingham, Wash.) 6 (2): 024008.

4. Berenguer, Roberto, María Del Rosario Pastor-Juan, Jesús Canales-Vázquez, Miguel Castro-García, María Victoria Villas, Francisco Mansilla Legorburo, and Sebastià Sabater. 2018. “Radiomics of CT Features May Be Nonreproducible and Redundant: Influence of CT Acquisition Parameters.” Radiology 288 (2): 407–15.

5. Boughdad, Sarah, Christophe Nioche, Fanny Orlhac, Laurine Jehl, Laurence Champion, and Irène Buvat. 2018. “Influence of Age on Radiomic Features in 18F-FDG PET in Normal Breast Tissue and in Breast Cancer Tumors.” Oncotarget 9 (56): 30855–68.

6. Choe, Jooae, Sang Min Lee, Kyung-Hyun Do, Gaeun Lee, June-Goo Lee, Sang Min Lee, and Joon Beom Seo. 2019. “Deep Learning-Based Image Conversion of CT Reconstruction Kernels Improves Radiomics Reproducibility for Pulmonary Nodules or Masses.” Radiology 292 (2): 365–73.

7. Duda, Dorota, Marek Kretowski, and Johanne Bezy-Wendling. 2013. “Effect of Slice Thickness on Texture-Based Classification of Liver Dynamic CT Scans.” In Computer Information Systems and Industrial Management, 96–107. Springer Berlin Heidelberg.

8. Du, Qian, Michael Baine, Kyle Bavitz, Josiah McAllister, Xiaoying Liang, Hongfeng Yu, Jeffrey Ryckman, et al. 2019. “Radiomic Feature Stability across 4D Respiratory Phases and Its Impact on Lung Tumor Prognosis Prediction.” PloS One 14 (5): e0216480.

9. Fave, Xenia, Dennis Mackin, Jinzhong Yang, Joy Zhang, David Fried, Peter Balter, David Followill, et al. 2015. “Can Radiomics Features Be Reproducibly Measured from CBCT Images for Patients with Non-Small Cell Lung Cancer?” Medical Physics 42 (12): 6784–97.

10. Fornacon-Wood, Isabella, Hitesh Mistry, Christoph J. Ackermann, Fiona Blackhall, Andrew McPartlin, Corinne Faivre-Finn, Gareth J. Price, and James P. B. O’Connor. 2020. “Reliability and Prognostic Value of Radiomic Features Are Highly Dependent on Choice of Feature Extraction Platform.” European Radiology. https://doi.org/10.1007/s00330-020-06957-9.

11. Griethuysen, Joost J. M. van, Andriy Fedorov, Chintan Parmar, Ahmed Hosny, Nicole Aucoin, Vivek Narayan, Regina G. H. Beets-Tan, Jean-Christophe Fillion-Robin, Steve Pieper, and Hugo J. W. L. Aerts. 2017. “Computational Radiomics System to Decode the Radiographic Phenotype.” Cancer Research 77 (21): e104–7.

12. Hosny, Ahmed, Chintan Parmar, John Quackenbush, Lawrence H. Schwartz, and Hugo J. W. L. Aerts. 2018. “Artificial Intelligence in Radiology.” Nature Reviews. Cancer 18 (8): 500–510.

13. Hu, Panpan, Jiazhou Wang, Haoyu Zhong, Zhen Zhou, Lijun Shen, Weigang Hu, and Zhen Zhang. 2016. “Reproducibility with Repeat CT in Radiomics Study for Rectal Cancer.” Oncotarget. https://doi.org/10.18632/oncotarget.12199.

14. Huynh, Elizabeth, Thibaud P. Coroller, Vivek Narayan, Vishesh Agrawal, John Romano, Idalid Franco, Chintan Parmar, Ying Hou, Raymond H. Mak, and Hugo J. W. L. Aerts. 2017. “Associations of Radiomic Data Extracted from Static and Respiratory-Gated CT Scans with Disease Recurrence in Lung Cancer Patients Treated with SBRT.” PloS One 12 (1): e0169172.

15. Kakino, Ryo, Mitsuhiro Nakamura, Takamasa Mitsuyoshi, Takashi Shintani, Hideaki Hirashima, Yukinori Matsuo, and Takashi Mizowaki. 2020. “Comparison of Radiomic Features in Diagnostic CT Images with and without Contrast Enhancement in the Delayed Phase for NSCLC Patients.” Physica Medica: PM: An International Journal Devoted to the Applications of Physics to Medicine and Biology: Official Journal of the Italian Association of Biomedical Physics 69 (January): 176–82.

16. Kim, Hyungjin, Chang Min Park, Myunghee Lee, Sang Joon Park, Yong Sub Song, Jong Hyuk Lee, Eui Jin Hwang, and Jin Mo Goo. 2016. “Impact of Reconstruction Algorithms on CT Radiomic Features of Pulmonary Tumors: Analysis of Intra- and Inter-Reader Variability and Inter-Reconstruction Algorithm Variability.” PloS One 11 (10): e0164924.

17. Kim, Young Jae, Hyun-Ju Lee, Kwang Gi Kim, and Seung Hyun Lee. 2019. “The Effect of CT Scan Parameters on the Measurement of CT Radiomic Features: A Lung Nodule Phantom Study.” Computational and Mathematical Methods in Medicine 2019 (February): 8790694.

18. Kolossváry, Márton, Bálint Szilveszter, Júlia Karády, Zsófia Dóra Drobni, Béla Merkely, and Pál Maurovich-Horvat. 2018. “Effect of Image Reconstruction Algorithms on Volumetric and Radiomic Parameters of Coronary Plaques.” Journal of Cardiovascular Computed Tomography, November. https://doi.org/10.1016/j.jcct.2018.11.004.

19. Kumar, Virendra, Yuhua Gu, Satrajit Basu, Anders Berglund, Steven A. Eschrich, Matthew B. Schabath, Kenneth Forster, et al. 2012. “Radiomics: The Process and the Challenges.” Magnetic Resonance Imaging 30 (9): 1234–48.

20. Lafata, Kyle, Jing Cai, Chunhao Wang, Julian Hong, Chris R. Kelsey, and Fang-Fang Yin. 2018. “Spatial-Temporal Variability of Radiomic Features and Its Effect on the Classification of Lung Cancer Histology.” Physics in Medicine and Biology 63 (22): 225003.

21. Larue, Ruben T. H. M., Janna E. van Timmeren, Evelyn E. C. de Jong, Giacomo Feliciani, Ralph T. H. Leijenaar, Wendy M. J. Schreurs, Meindert N. Sosef, et al. 2017. “Influence of Gray Level Discretization on Radiomic Feature Stability for Different CT Scanners, Tube Currents and Slice Thicknesses: A Comprehensive Phantom Study.” Acta Oncologica 56 (11): 1544–53.

22. Lee, Seung-Hak, Hwan-Ho Cho, Ho Yun Lee, and Hyunjin Park. 2019. “Clinical Impact of Variability on CT Radiomics and Suggestions for Suitable Feature Selection: A Focus on Lung Cancer.” Cancer Imaging: The Official Publication of the International Cancer Imaging Society 19 (1): 54.

23. Liguori, Carlo, Giulia Frauenfelder, Carlo Massaroni, Paola Saccomandi, Francesco Giurazza, Francesca Pitocco, Riccardo Marano, and Emiliano Schena. 2015. “Emerging Clinical Applications of Computed Tomography.” Medical Devices 8 (June): 265–78.

24. Liu, Zhenyu, Shuo Wang, D. Dong, Jingwei Wei, Cheng Fang, Xuezhi Zhou, Kai Sun, et al. 2019. “The Applications of Radiomics in Precision Diagnosis and Treatment of Oncology: Opportunities and Challenges.” Theranostics 9 (5): 1303–22.

25. Li, Yajun, Lin Lu, Manjun Xiao, Laurent Dercle, Yue Huang, Zishu Zhang, Lawrence H. Schwartz, Daiqiang Li, and Binsheng Zhao. 2018. “CT Slice Thickness and Convolution Kernel Affect Performance of a Radiomic Model for Predicting EGFR Status in Non-Small Cell Lung Cancer: A Preliminary Study.” Scientific Reports 8 (1): 17913.

26. Lo, P., S. Young, H. J. Kim, M. S. Brown, and M. F. McNitt-Gray. 2016. “Variability in CT Lung-Nodule Quantification: Effects of Dose Reduction and Reconstruction Methods on Density and Texture Based Features.” Medical Physics 43 (8): 4854.

27. Lu, Hong, Wei Mu, Yoganand Balagurunathan, Jin Qi, Mahmoud A. Abdalah, Alberto L. Garcia, Zhaoxiang Ye, Robert J. Gillies, and Matthew B. Schabath. 2019. “Multi-Window CT Based Radiomic Signatures in Differentiating Indolent versus Aggressive Lung Cancers in the National Lung Screening Trial: A Retrospective Study.” Cancer Imaging: The Official Publication of the International Cancer Imaging Society 19 (1): 45.

28. Lu, Lin, Ross C. Ehmke, Lawrence H. Schwartz, and Binsheng Zhao. 2016. “Assessing Agreement between Radiomic Features Computed for Multiple CT Imaging Settings.” PloS One 11 (12): e0166550.

29. Mackin, Dennis, Xenia Fave, Lifei Zhang, David Fried, Jinzhong Yang, Brian Taylor, Edgardo Rodriguez-Rivera, Cristina Dodge, Aaron Kyle Jones, and Laurence Court. 2015. “Measuring Computed Tomography Scanner Variability of Radiomics Features.” Investigative Radiology 50 (11): 757–65.

30. Mackin, Dennis, Rachel Ger, Cristina Dodge, Xenia Fave, Pai-Chun Chi, Lifei Zhang, Jinzhong Yang, et al. 2018. “Effect of Tube Current on Computed Tomography Radiomic Features.” Scientific Reports 8 (1): 2354.

31. Mahon, Rebecca Nichole, Geoffrey D. Hugo, and Elisabeth Weiss. 2019. “Repeatability of Texture Features Derived from Magnetic Resonance and Computed Tomography Imaging and Use in Predictive Models for Non-Small Cell Lung Cancer Outcome.” Physics in Medicine and Biology, April. https://doi.org/10.1088/1361-6560/ab18d3.

32. “MathWorks - Makers of MATLAB and Simulink.” n.d. accessed May 17, 2020. https://www.mathworks.com/.

33. Meyer, Mathias, James Ronald, Federica Vernuccio, Rendon C. Nelson, Juan Carlos Ramirez-Giraldo, Justin Solomon, Bhavik N. Patel, Ehsan Samei, and Daniele Marin. 2019. “Reproducibility of CT Radiomic Features within the Same Patient: Influence of Radiation Dose and CT Reconstruction Settings.” Radiology 293 (3): 583–91.

34. Midya, Abhishek, Jayasree Chakraborty, Mithat Gönen, Richard K. G. Do, and Amber L. Simpson. 2018. “Influence of CT Acquisition and Reconstruction Parameters on Radiomic Feature Reproducibility.” Journal of Medical Imaging (Bellingham, Wash.) 5 (1): 011020.

35. Moher, David, Larissa Shamseer, Mike Clarke, Davina Ghersi, Alessandro Liberati, Mark Petticrew, Paul Shekelle, Lesley A. Stewart, and PRISMA-P Group. 2015. “Preferred Reporting Items for Systematic Review and Meta-Analysis Protocols (PRISMA-P) 2015 Statement.” Systematic Reviews 4 (January): 1.

36. Ng, Francesca, Robert Kozarski, Balaji Ganeshan, and Vicky Goh. 2013. “Assessment of Tumor Heterogeneity by CT Texture Analysis: Can the Largest Cross-Sectional Area Be Used as an Alternative to Whole Tumor Analysis?” European Journal of Radiology 82 (2): 342–48.

37. Park, Ji Eun, Ho Sung Kim, Donghyun Kim, Seo Young Park, Jung Youn Kim, Se Jin Cho, and Jeong Hoon Kim. 2020. “A Systematic Review Reporting Quality of Radiomics Research in Neuro-Oncology: Toward Clinical Utility and Quality Improvement Using High-Dimensional Imaging Features.” BMC Cancer. https://doi.org/10.1186/s12885-019-6504-5.

38. Perrin, Thomas, Abhishek Midya, Rikiya Yamashita, Jayasree Chakraborty, Tome Saidon, William R. Jarnagin, Mithat Gonen, Amber L. Simpson, and Richard K. G. Do. 2018. “Short-Term Reproducibility of Radiomic Features in Liver Parenchyma and Liver Malignancies on Contrast-Enhanced CT Imaging.” Abdominal Radiology (New York) 43 (12): 3271–78.

39. Shafiq-Ul-Hassan, Muhammad, Kujtim Latifi, Geoffrey Zhang, Ghanim Ullah, Robert Gillies, and Eduardo Moros. 2018. “Voxel Size and Gray Level Normalization of CT Radiomic Features in Lung Cancer.” Scientific Reports 8 (1): 10545.

40. Shafiq-ul-Hassan, Muhammad, Geoffrey G. Zhang, Dylan C. Hunt, Kujtim Latifi, Ghanim Ullah, Robert J. Gillies, and Eduardo G. Moros. 2017. “Accounting for Reconstruction Kernel-Induced Variability in CT Radiomic Features Using Noise Power Spectra.” The Journal of Medical Investigation: JMI 5 (1): 011013.

41. Shafiq-Ul-Hassan, Muhammad, Geoffrey G. Zhang, Kujtim Latifi, Ghanim Ullah, Dylan C. Hunt, Yoganand Balagurunathan, Mahmoud Abrahem Abdalah, et al. 2017. “Intrinsic Dependencies of CT Radiomic Features on Voxel Size and Number of Gray Levels.” Medical Physics 44 (3): 1050–62.

42. Solomon, Justin, Achille Mileto, Rendon C. Nelson, Kingshuk Roy Choudhury, and Ehsan Samei. 2016. “Quantitative Features of Liver Lesions, Lung Nodules, and Renal Stones at Multi-Detector Row CT Examinations: Dependency on Radiation Dose and Reconstruction Algorithm.” Radiology 279 (1): 185–94.

43. Traverso, Alberto, Leonard Wee, Andre Dekker, and Robert Gillies. 2018. “Repeatability and Reproducibility of Radiomic Features: A Systematic Review.” International Journal of Radiation Oncology*Biology*Physics. https://doi.org/10.1016/j.ijrobp.2018.05.053.

44. Varghese, Bino A., Darryl Hwang, Steven Y. Cen, Joshua Levy, Derek Liu, Christopher Lau, Marielena Rivas, Bhushan Desai, David J. Goodenough, and Vinay A. Duddalwar. 2019. “Reliability of CT-Based Texture Features: Phantom Study.” Journal of Applied Clinical Medical Physics / American College of Medical Physics 20 (8): 155–63.

45. Yamashita, Rikiya, Thomas Perrin, Jayasree Chakraborty, Joanne F. Chou, Natally Horvat, Maura A. Koszalka, Abhishek Midya, et al. 2019. “Radiomic Feature Reproducibility in Contrast-Enhanced CT of the Pancreas Is Affected by Variabilities in Scan Parameters and Manual Segmentation.” European Radiology, August. https://doi.org/10.1007/s00330-019-06381-8.

46. Yasaka, Koichiro, Hiroyuki Akai, Dennis Mackin, Laurence Court, Eduardo Moros, Kuni Ohtomo, and Shigeru Kiryu. 2017. “Precision of Quantitative Computed Tomography Texture Analysis Using Image Filtering: A Phantom Study for Scanner Variability.” Medicine 96 (21): e6993.

47. Zhang, Lifei, David V. Fried, Xenia J. Fave, Luke A. Hunter, Jinzhong Yang, and Laurence E. Court. 2015. “IBEX: An Open Infrastructure Software Platform to Facilitate Collaborative Work in Radiomics.” Medical Physics 42 (3): 1341–53.

48. Zhao, Binsheng, Yongqiang Tan, Wei-Yann Tsai, Jing Qi, Chuanmiao Xie, Lin Lu, and Lawrence H. Schwartz. 2016. “Reproducibility of Radiomics for Deciphering Tumor Phenotype with Imaging.” Scientific Reports 6 (March): 23428.

